# Genetic modifiers of rare variants in monogenic developmental disorder loci

**DOI:** 10.1101/2022.12.15.22283523

**Authors:** Rebecca Kingdom, Robin N. Beaumont, Andrew R. Wood, Michael N. Weedon, Caroline F. Wright

## Abstract

Rare damaging variants in a large number of genes are known to cause monogenic developmental disorders (DD), and have been shown to cause milder sub-clinical phenotypes in population cohorts. To investigate potential genetic modifiers, we identified individuals in UK Biobank with predicted deleterious variants in 599 autosomal dominant DD genes, and found that carrying multiple rare variants in these genes had an additive adverse effect on numerous cognitive and socio-economic traits, which could be partially counterbalanced by a higher educational attainment polygenic score (EA-PGS). Amongst rare DD variant carriers, those with a DD-related clinical diagnosis had a substantially lower EA-PGS and more severe phenotype than those without. Our results suggest that the overall burden of both rare and common variants can modify the expressivity of a phenotype, which may influence whether an individual reaches the threshold for clinical disease.

## INTRODUCTION

Ascertaining whether rare genetic variants cause a monogenic phenotype can be challenging due to incomplete penetrance and variable expressivity^1^. Many rare variant studies use clinical or familial cohorts that can overestimate the penetrance of causal variants^2^. The presence of such rare, putatively damaging variants in healthy population cohorts^3^ can provide a lower boundary for estimates of penetrance, and individuals in both clinical and population cohorts display a spectrum of phenotypic variability caused by similar or identical variants^1,4^. Previous research has suggested that common genetic variants can modify the penetrance or expressivity of phenotypes caused by rare genetic variants^5–7^, potentially through the liability threshold model, which posits that a certain threshold of disease susceptibility needs to be crossed before clinically-diagnosable disease manifests^8–11^. Some damaging rare variants may reach this threshold alone, resulting in a monogenic disease phenotype with 100% penetrance, while other variants may need additional genetic, environmental, or other modifiers to reach this threshold^8^. In certain diseases, common variant burden has been shown to confer a risk similar to that of a deleterious monogenic variant, where the highest polygenic risk may be equivalent to that conferred by a monogenic variant^12,13^. As the effect of each individual common variant is very small^14^, aggregating them together as a polygenic score (PGS) has become a widely used method for predicting overall risk^15,16^, and combining PGS with rare pathogenic variants could improve individual disease prediction^17,18^.

Previously, we showed that rare, predicted loss-of-function (pLoF), deleterious missense and large copy number variants (CNVs) in genes and loci linked with severe monogenic developmental disorders (DD) can have milder, sub-clinical effects in the general population^19^. Related common variant burden has been shown to affect the phenotype in carriers of such variants^5,20^, suggesting that the cumulative effect of common variants can modify the penetrance of rare variants in such phenotypes even if the primary cause is thought to be monogenic. While the impact of common variants on overall phenotypic expressivity has been examined for several neuropsychiatric^21–23^ and other disease cohorts^24–26^, the modification of rare variant penetrance by other rare genetic variants has not been widely investigated due to the very large cohort sizes required. Here, we present an analysis of common and rare variant burden in 419,854 adults from the UK Biobank (UKB)^27^. We investigate individuals carrying a rare pLoF variant in genes and loci where similar variants are known to cause monogenic DD, and use related polygenic scores and additional rare variant burden to examine the effect on a number of related cognitive phenotypes and socioeconomic traits. We show that rare variant burden across these loci and PGS for Educational Attainment (EA-PGS) has an additive effect on the phenotype. Our results demonstrate that both rare and common genetic variants linked to relevant traits can contribute towards the variable expressivity of rare, predicted large-effect variants in known monogenic disease.

## RESULTS

We used exome sequencing and microarray data from individuals in UKB who were of genetically defined European ancestry (N = 419,854). We identified carriers of rare (allele count =<5) pLoF^28^ or deleterious missense (REVEL>0.7)^29^ variants in any of 599 genes from the Developmental Disorders Gene-2-Phenotype Database (DDG2P)^19,30^ in which damaging rare variants are a known cause of autosomal dominant DD. Carriers of multigenic CNVs were also included where the variant overlapped known syndromic DD-related loci^31,32^, as described previously^19^. We calculated a published EA-PGS^33^ using summary statistics and weighted allele effects from genome-wide association studies (GWAS) for every individual in UKB with European ancestry. Phenotypes of interest were selected from self-reported questionnaires, based on their relevance to cognitive, behavioural, reproductive, and socio-economic traits related to neurodevelopmental disorders **(Supplementary Table 1)**. In addition, clinically relevant diagnoses were identified using ICD-9 or ICD-10 codes from Hospital Episode Statistics and combined into three categories: child-DD, adult-DD (schizophrenia or bipolar disorder), and other mental health issues (neurotic and anxiety disorders; see **Supplementary Table 2)**.

We first investigated whether DD-related phenotypes could be modified amongst rare DD variant carriers by the presence of additional rare pLoF or damaging missense variants in the same set of DDG2P genes. In UKB, 50,395 (12%) individuals carry a single rare likely deleterious variant overlapping one of the 599 autosomal dominant DDG2P genes (12,153 pLoF and 35,603 missense) or syndromic DD loci (1127 large deletions and 1512 large duplications); an additional 3831 individuals carry two rare DD variants, and 219 individuals have three or more putatively deleterious rare variants across these DD loci. The highest overall rare variant burden across the DD loci was five, which was seen in two individuals with three missense variants and two pLoF variants each (**Supplementary Table 3**). We performed regression analysis to test associations between number of rare variants in DD genes and 15 DD-related traits and diagnoses, using linear regression for continuous traits **(Figure 1a)** and logistic regression for binary traits **(Figure 1b)**. Increasing rare variant burden correlated with a larger change away from the average UKB participant in several DD-related phenotypes, including lower fluid intelligence, shorter stature, lower income, lower likelihood of being employed, lower likelihood of being a parent, and higher Townsend Deprivation Index (TDI). An increase in rare variant burden also correlated with a higher likelihood of having a DD-related diagnosis, and those with three or more rare DD variants were 2.1X (95% CI: 1.05-4.33, p = 0.03) and 1.7X (95% CI: 1.01=2.89, p = 0.04) more likely to be diagnosed with a child-DD or an adult DD-related diagnosis respectively than non-carriers **(Figure 1b)**. When we excluded those with rare missense variants and only considered pLoF and large CNV carriers, we observed a larger change in phenotype, but the smaller number of individuals present in each group reduced the statistical power substantially; nonetheless, those with two or three rare variants were 2.2X (95% CI: 1.37-3.43, p = 0.0009) more likely to have a child DD related diagnosis than those without a pLoF variant or CNV **(Supplementary Table 4)**.

**Figure 1:**
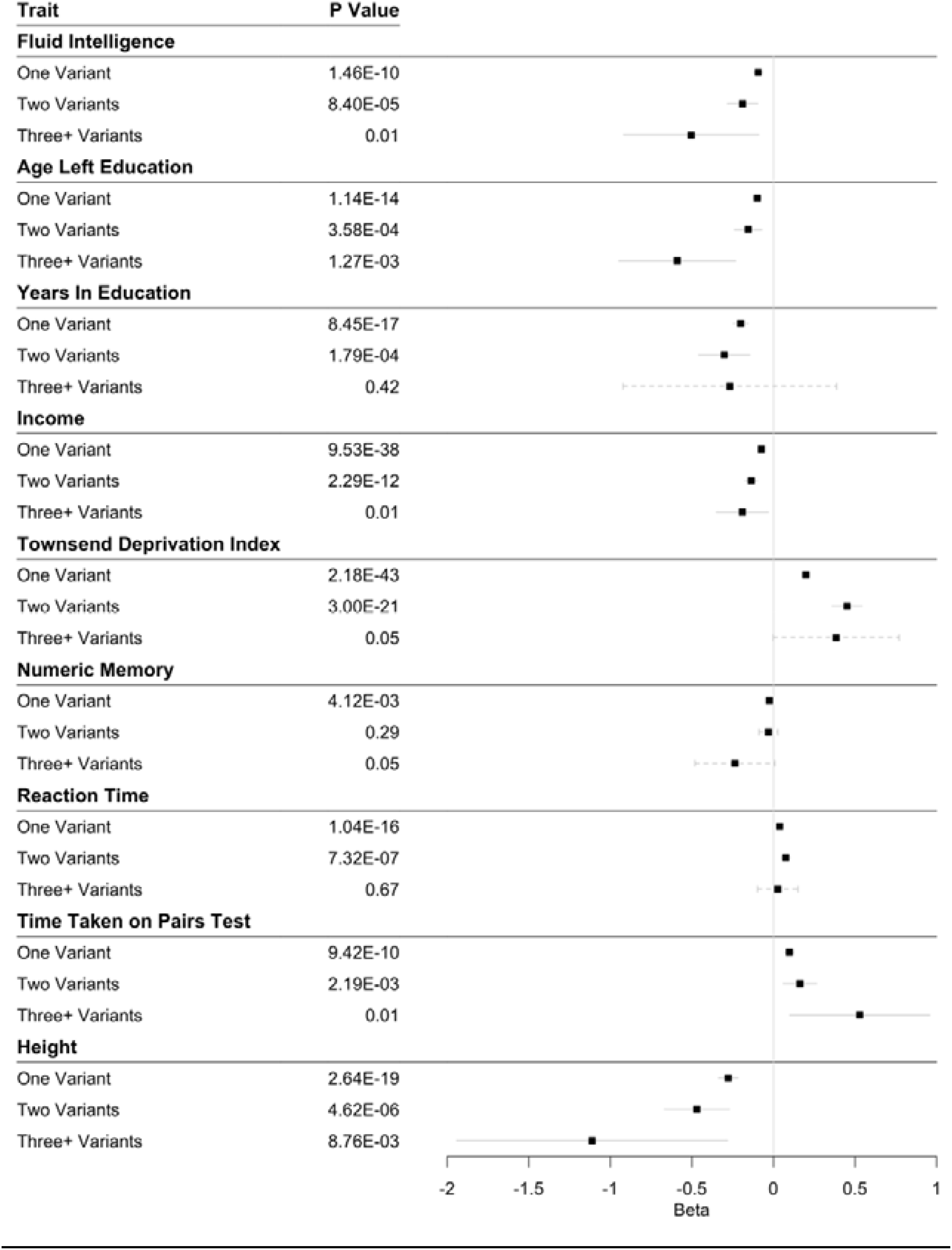
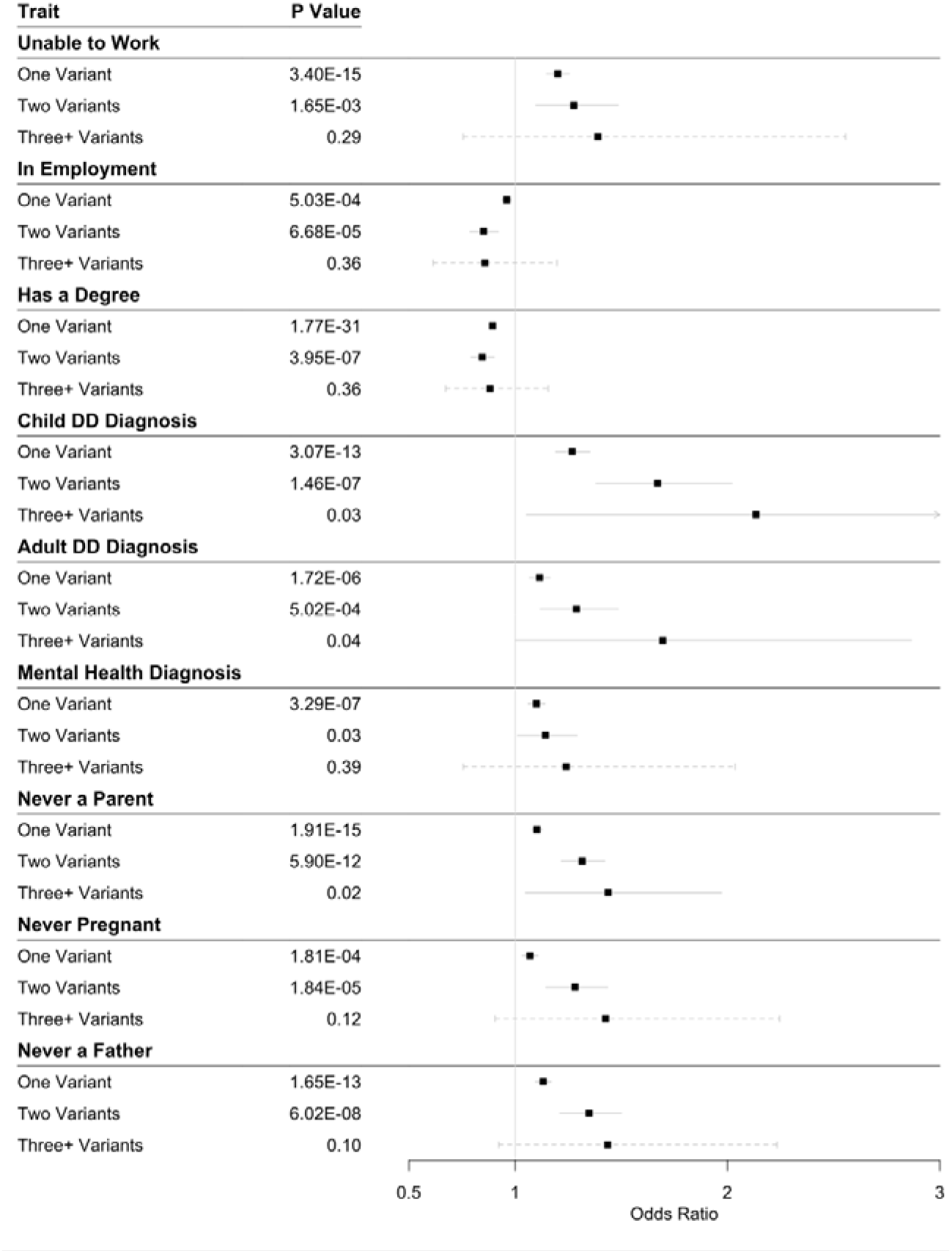
Effect of rare DD variant burden on DD-related phenotypes in UKB. Associations of **(a)** continuous traits and **(b)** binary traits/diagnoses in individuals carrying either 1, 2 or 3+ rare pLoF, deleterious missense, or multigenic variants overlapping dominant DDG2P genes, compared with the rest UKB (i.e. non-carriers). Beta values for continuous traits were measured as follows: Fluid Intelligence = standardised units (ranging from 1-13); Age Left Education and Years in Education = years; Height = cm; Reaction Time, Time taken on Pairs Test, Numeric Memory, Income, and Townsend Deprivation Index (TDI) = standard deviations from the mean.

Next, we investigated the effect of common polygenic background on rare DD variant carriers^9^. We separated the UKB cohort into five EA-PGS quantiles and repeated the phenotype association tests with rare DD variant carrier status. We saw a similar trend across all traits tested against the EA-PGS quintiles (**Supplementary Figure 1**), with the direction of the PGS effect being the same in both carrier and non-carrier groups. Individuals who carried at least one rare variant showed a consistently larger change in fluid intelligence, years of education, employment and TDI across the PGS spectrum compared to the control group, with larger phenotypic effects observed in carriers of multiple rare DD variants **(Figure 2)**. We observed similar trends when we repeated this analysis excluding missense variants (**Supplementary Table 5)** or using a smaller subset of DD genes **(Supplementary Table 6)**, specifically those known to cause disease via haploinsufficiency (n= 325) or only those that reached genome-wide significance based on burden of *de novo* variants in ∼31,000 DD cases (n=125)^34^.

**Figure 2:**
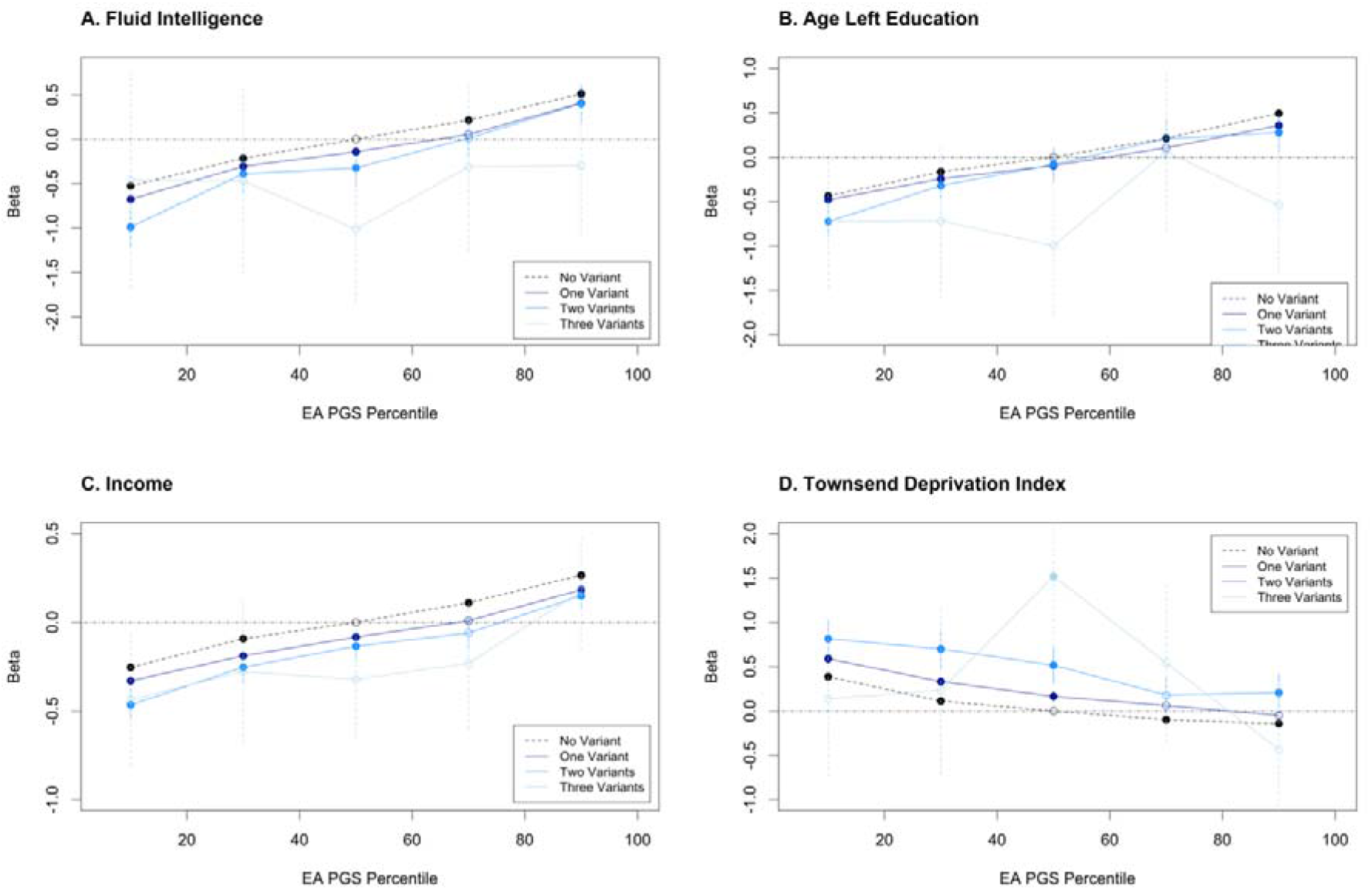
Additive effect of rare DD variant burden and EA-PGS on DD-related phenotypes. Change in **(a)** fluid intelligence, **(b)** income, **(c)** years in education and **(d)** Townsend Deprivation Index (TDI) are shown versus EA-PGS quintile in UKB. Black dashed line = non-carriers of rare DD variants (N=365,409); dark/medium/light blue lines = carriers of 1, 2 or 3+ rare DD variants (N= 50,395, 3831, and 219 respectively). Notably, within UKB, a high enough EA-PGS can compensate for the presence of a primary variant and, in most cases, any additional rare DD variants.

For fluid intelligence, the difference in the mean score between the bottom and top EA-PGS quintiles equated to approximately 1 point on the 13-point scale, both for rare variant carriers and non-carriers in UKB. Rare DD variant carrier status was equivalent to around a 20-percentile point decrease in EA-PGS, on average, with the result that an EA-PGS above the 70^th^ centile was able to compensate for the effect of carrying a single rare DD variant on fluid intelligence **(Supplementary Table 7)**. Importantly, rare variant carrier status and EA-PGS appear to have an additive effect when assessed against multiple related traits, with the effect of rare variants remaining similar throughout the EA-PGS spectrum. When we investigated rare variant classes within fluid intelligence scores, deleterious missense variant carriers reached parity with the control group at the 62^nd^ EA-PGS percentile, LoF carriers at the 80^th^ percentile and CNV duplication carriers at the 82^nd^ percentile, while CNV deletion carriers never reached parity with the control group **(Supplementary Table 7)**. We hypothesized that the EA-PGS could include SNPs in cis-regulatory regions of monogenic DDG2P genes, so we examined proximity between the 599 autosomal dominant DDG2P genes and 3952 SNPs included in the EA-PGS, using simulations to test whether the genes fall disproportionately close to the GWAS loci^35^. As expected, we found that found that the GWAS loci were closer to DDG2P genes than expected by chance (p = 0.005), suggesting that the large-effect rare variants and small-effect common variants may work through overlapping biological pathways.

As the UKB cohort is known to be biased towards healthier, wealthier, and more educated individuals than the general population^36^, we hypothesized that those individuals in UKB who carry a rare DD variant might also have a higher EA-PGS on average than the non-carrier control group, which partially compensates for the potentially deleterious effects of the rare DD variant. Overall, we observed that individuals who carried at least one rare DD variant did indeed have a slightly higher EA-PGS percentile than non-carriers (t-test: difference =+2.1, 95%CI: 1.9-2.4, p < 0.0005), supporting this hypothesis. Furthermore, among the small number of individuals who achieved the top score on the fluid intelligence test (N=139), we observed that rare DD variant carriers (N=4) were depleted versus the rest of UKB (3% versus 13%, p = 0.0002) and had a substantially higher EA-PGS percentile than non-carriers (t-test: difference = +26.1, 95%CI: 1.8-50.3, p = 0.04).

Intrigued by the presence of these apparently highly intelligent rare DD variant carriers, we further investigated phenotypic “deviators” in whom the predicted genetic susceptibility is discordant with the observed phenotype^37^, e.g. high EA-PGS but low fluid intelligence score and *vice versa* **(Figure 3a)**. This question has particular clinical relevance, as it has previously been suggested that individuals with familial disease could be prioritised for genetic testing based on having a low-risk PGS, as they may be more likely to have a single large-effect causal variant than individuals with a high-risk PGS whose disease may be more polygenic^38,39^. To investigate this hypothesis, we further split the UKB cohort into deciles of EA-PGS and tested whether individuals whose low cognitive phenotype was discordant with their high EA-PGS were more likely to be rare DD variant carriers than the remainder of the UKB cohort. Individuals in the top EA-PGS decile but with low fluid intelligence (scores of 0 or 1 out of 13) were more likely to be rare DD variant carriers (OR: 1.68, 95% CI: 1.13-2.50, p = 0.01) **(Figure 3b)**, when compared to those in the same EA-PGS decile who did not have a low fluid intelligence score, as were those in the top EA-PGS decile who had no educational qualifications on record (OR: 1.22, 95% CI: 1.10-1.35, p = 0.00006) **(Figure 3c)**. When separated by rare DD variant class, we found that large multigenic deletions had a larger effect than any other type of rare DD variant (OR: 4.7, 95% CI: 1.73-12.95, p = 0.002), followed by pLoF variants, and then CNV duplications **(Supplementary Table 8)**. We then investigated whether the opposite was also true, i.e. whether those with a bottom decile EA-PGS but a high fluid intelligence score (11-13 out of 13) were less likely to be rare variant carriers, and found that individuals were almost half as likely as others in the same decile to carry a rare DD variant (OR: 0.58, 95% CI: 0.38-0.87, p = 0.009).

**Figure 3:**
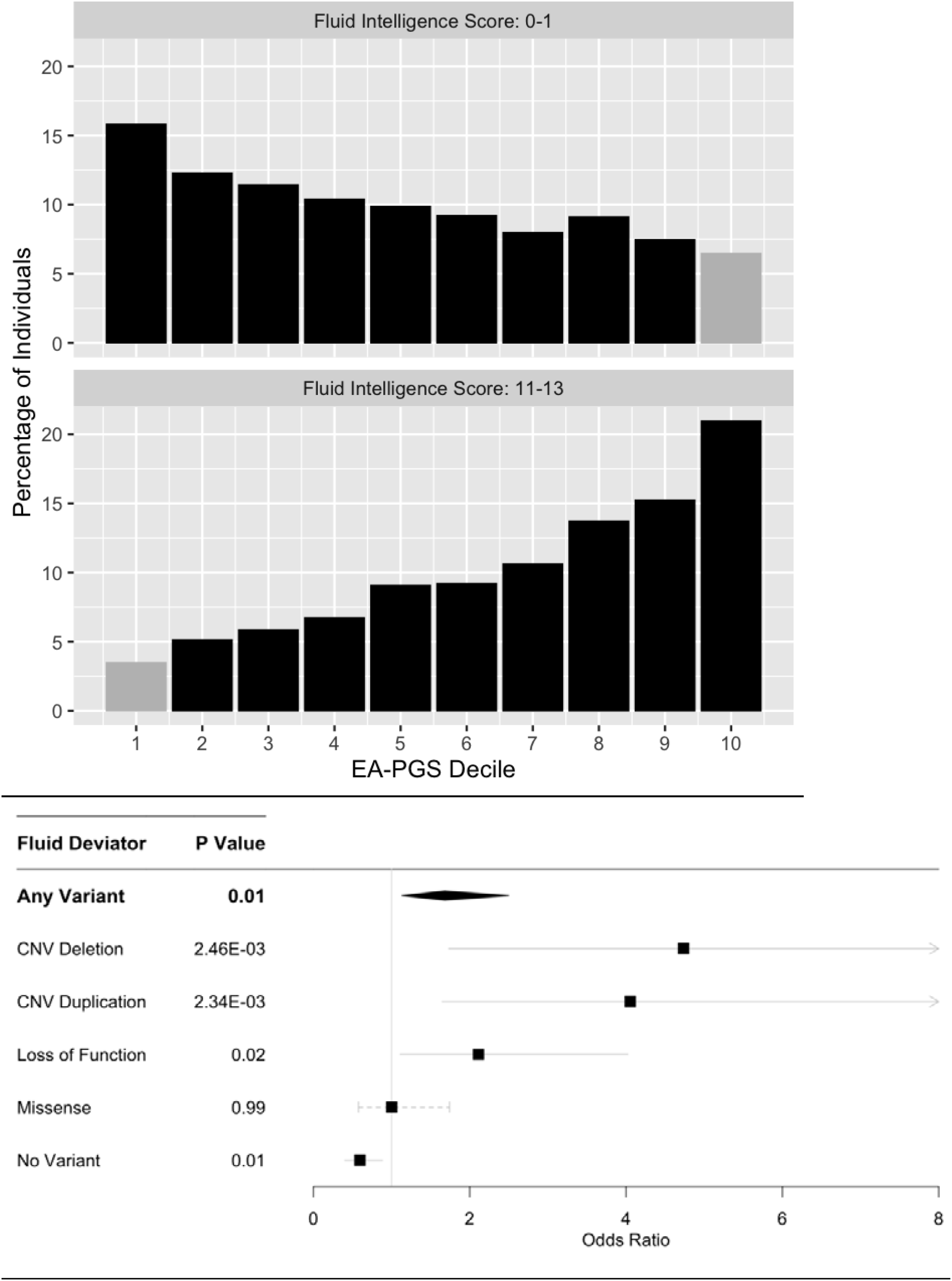
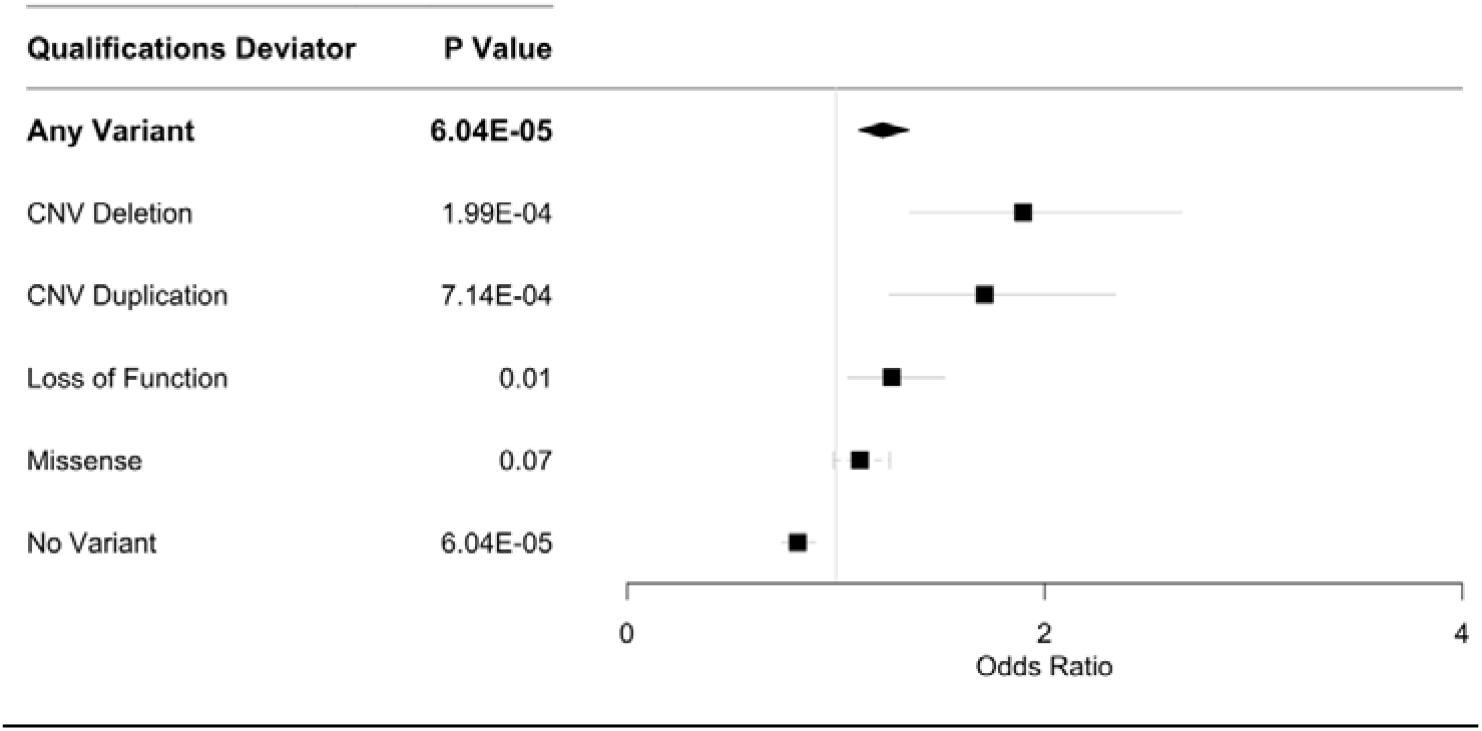
Rare DD variant carrier status of phenotypic “deviators” from EA-PGS predictions. **(a)** Distribution of EA-PGS and fluid intelligence within UKB; phenotypic “deviators” are highlighted and defined as either a top decile EA-PGS and a low fluid intelligence score (0 or 1) or a bottom decile EA-PGS and high fluid intelligence score (11, 12 or 13). **(b)** Individuals in UKB who have a top decile EA-PGS but scored low on the fluid intelligence test or **(c)** reported having no qualifications recorded despite having a top decile EA-PGS were more likely to be rare DD variant carriers. The comparator group is those within the same EA-PGS but a higher fluid intelligence score, or recorded qualifications.

Finally, we investigated whether a decrease in EA-PGS correlates with the likelihood of receiving a clinical diagnosis related to DD amongst the rare DD variant carriers we identified in UKB. The number of individuals identified within the three diagnostic categories (child-DD N=7933; adult-DD N=19,004; and other mental health issues N=32,911) is likely to be under-estimated due to absence of, or omissions in, individual hospital records available within UKB. Therefore, while individuals in any of these diagnostic categories were more likely to be rare DD variant carriers than the rest of UKB, the majority did not carry a rare variant in any of the DD genes, and many individuals with a rare DD variant did not have a corresponding diagnosis. Despite these limitations, we found that, amongst rare DD variant carriers, those with a related clinical diagnosis across any of our three categories had a substantially lower EA-PGS than those without **(Figure 4)**. They also had a larger phenotypic change than other rare variant carriers without a diagnosis; individuals with a rare DD variant and a related clinical diagnosis were more likely to be unable to work (OR: 6.66, 95% CI: 6.07-7.32, p = 4.51E-308), less likely to have a degree (OR: 0.71, 95% CI: 0.66-0.76, p = 3.76E-23), and less likely to be in employment (OR: 0.33, 95% CI: 0.31-0.37, p = 2.07E-143) than those who carry a rare DD variant but do not have a diagnosis recorded in UKB **(Supplementary Table 9)**. This suggests that both the aggregation of overall number of rare DD variants carried and a lower EA-PGS can alter the overall expressivity of the phenotype towards reaching the threshold of clinical disease.

**Figure 4:**
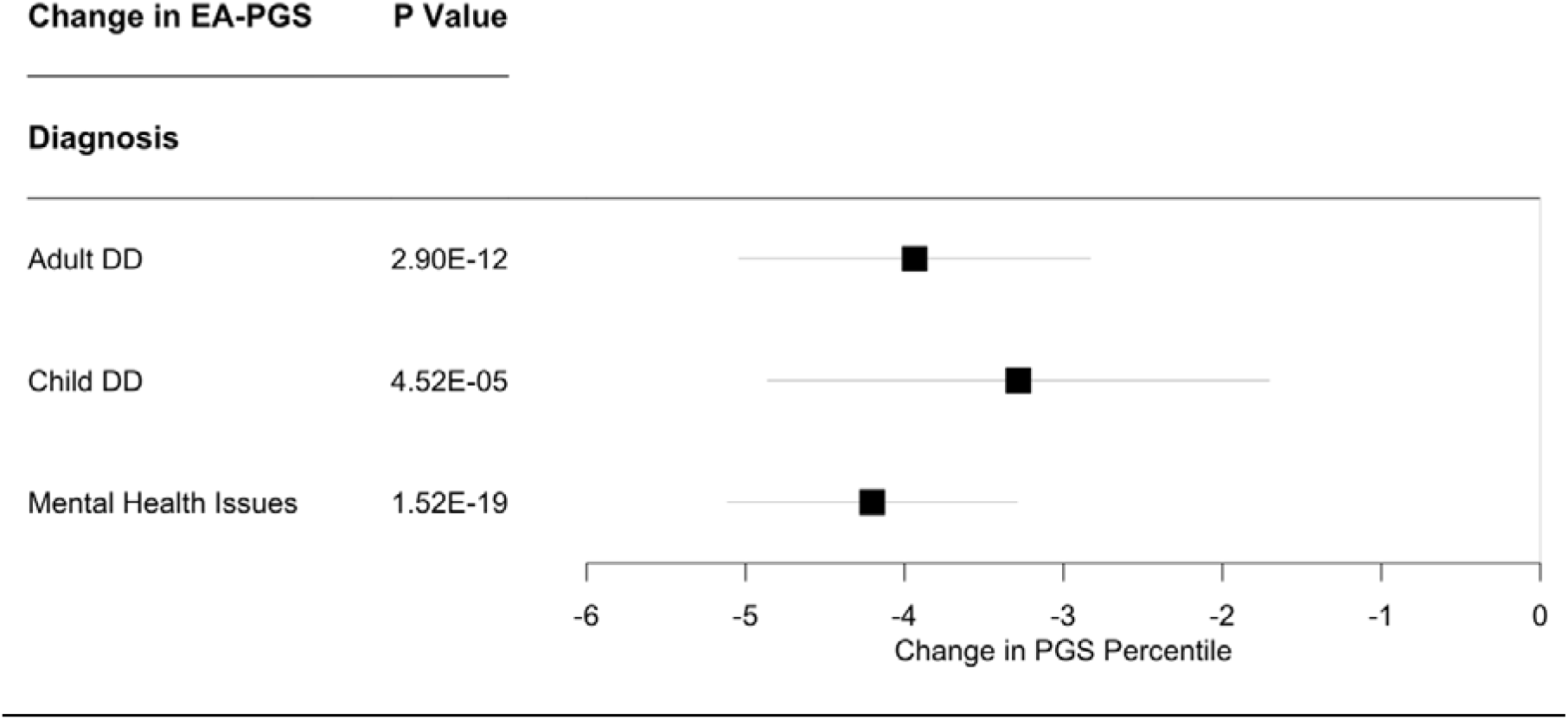
Average change in EA-PGS among rare DD variant carriers with a relevant clinical diagnosis. Amongst individuals carrying one or more rare DD variants, those who are clinically diagnosed with either child-DD or adult-DD or other mental health issues have substantially a lower EA-PGS than those who do not have a related clinical diagnosis recorded in UKB.

## DISCUSSION

We have shown that the phenotypic effect of rare and common genetic variants is additive for a genetically heterogeneous rare disease in a population cohort. The adverse effects of carrying a single deleterious rare variant in genes wherein similar variants cause monogenic DD can be modified by additional rare variants in those genes or by common variants across the genome. Carriers of multiple rare DD variants in UKB have lower fluid intelligence, shorter stature, fewer children, lower income, higher unemployment and higher TDI compared with carriers of single rare DD variants. Additionally, our results suggest that having a higher EA-PGS can partially compensate for the negative cognitive and socio-economic effects of carrying a single or multiple rare DD variants. Moreover, a higher burden of DD-associated variants is more likely to push the phenotypic presentation over the threshold for clinical diagnosis, and correlates with a larger change in phenotype compared to individuals who carry fewer or no variants. Our results suggest that PGS may provide some clinical utility by improving diagnostic interpretation of rare, likely pathogenic variants that cause monogenic disease.

Investigating the effect of pathogenic rare variants in the general population is important for understanding penetrance and variable expressivity of monogenic diseases. However, there are important limitations on using large-scale genetic data from UK Biobank to investigate rare disease. Firstly, some of the deleterious rare variants we identified may be benign, due to technical artefacts, erroneous pathogenicity predictions, alternative splicing or other mechanisms. Secondly, UKB is known to have an ascertainment bias towards healthier and wealthier individuals compared with the rest of the British population^36^, and individuals affected by severe highly penetrant monogenic disorders are likely to be depleted from the cohort. Thirdly, complete medical histories are not available within UKB, which is a relatively old cohort, so many phenotypes of relevance to childhood DDs cannot be evaluated. Fourthly, environment influences were not assessed and may have additional effects on the overall phenotype^40,41^ as well as altering the penetrance and expressivity of genetic variants through gene-environment interactions. Finally, there are challenges in applying common variant PGS across a population, as the underlying summary statistics are heavily dependent upon the populations and ethnicities in which the GWAS were performed. Moreover, PGS often include GWAS results from meta-analyses that incorporate the UKB cohort, which could result in some overfitting. Nonetheless, despite these limitations, our results are consistent with previous studies showing the effect of rare DD variants in non-clinical cohorts and the modifying effect of PGS on carriers of rare DD variants^5,6^.

In conclusion, we have shown that common and rare genetic variants can additively and independently affect the phenotype of non-clinically ascertained individuals. Our results go some way to explaining the puzzling observation of apparently healthy carriers of monogenic disease-causing variants in the general population, as well as instances of incomplete penetrance and variable expressivity in families affected by rare diseases. Further research is needed to investigate other modifiers, such as rare non-coding variants and gene-environment interactions, and to understand the mechanisms by which genetic modifiers act. Ultimately, incorporating the additive effects of both rare and common variants will improve our understanding of disease.

## METHODS

### UK Biobank Cohort

UKB is a voluntary population-based cohort from the UK with deep phenotyping data and genetic data for around 500,000 individuals aged 40-70 years at recruitment. Individuals provided a variety of information via self-report questionnaires, cognitive and anthropometric measurements, and Hospital Episode Statistics (HES) including ICD-9 and ICD-10 codes. Genotypes for single nucleotide polymorphisms (SNPs) were generated using the Affymetrix Axiom UK Biobank array (∼450,000 individuals) and the UK BiLEVE array (∼50,000 individuals). This dataset underwent extensive central quality control (http://biobank.ctsu.ox.ac.uk). A subset of ∼450,000 individuals also underwent exome sequencing using the IDT xGen Exome Research Panel v1.0; this dataset was made available for research in October 2021. Detailed sequencing and variant detection methodology for UKB is available at https://biobank.ctsu.ox.ac.uk/showcase/label.cgi?id=170. The UKB resource was approved by the UK Biobank Research Ethics Committee and all participants provided written informed consent to participate.

### Gene selection

We used the clinically curated Developmental Disorders Gene2Phenotype Database (DDG2P) to select genes known to cause monogenic DD. The database (accessed from https://www.ebi.ac.uk/gene2phenotype/ on 27 November 2020) was constructed and clinically curated from published literature and provides information relating to genes, variants and phenotypes associated with DDs, including mode of inheritance and mechanism of pathogenicity. We included all genes that had been annotated as monoallelic (i.e., autosomal dominant) with an evidence level of “confirmed” or “probable” (n=599).

### Variant selection

We used exome sequencing data from 419,854 individuals in UKB to identify carriers of rare, single nucleotide variants (SNVs) and/or insertions/deletions (indels) in any of the selected DDG2P genes. For our analyses, rare was defined as any variant that occurred in 5 or fewer individuals in the UKB cohort. We selected two functional classes of variant in canonical transcripts based on annotation by the Ensembl Variant Effect Predictor (v104)^30^: (1) likely deleterious LoF variants: we defined a LoF variant as one that is predicted to cause a premature stop, a frameshift, or abolish a canonical splice site; only those variants deemed to be high confidence by the Loss-Of-Function Transcript Effect Estimator (LOFTEE) were retained (https://github.com/konradjk/loftee); and (2) likely deleterious missense variants: missense variants with a REVEL score > 0.7. Individuals with >1 variant within a 40bp window in the same gene were counted once. In addition, we used SNP-array data from 488,377 genotyped individuals in UKB and PennCNV^42^ (version 1.0.4) to detect multigenic CNVs overlapping 69 published CNVs strongly associated with developmental delay, as described previously^19^.

### PGS calculation

We created the educational attainment PGS using GWAS summary statistics from a large cohort meta-analysis, using 3952 SNPs for the Educational Attainment (EA) PGS, with data from Okbay *et al*. 2022^33^. The EA-PGS was calculated as ∑_i_w_i_g_i_, where w is the weight (effect size) for SNP I and g_i_ is the genotype (number of effect alleles, 0-2) at SNP i. The SNP weightings were the regression coefficients obtained from the most recently reported GWAS as mentioned above.

### Statistical analysis

We performed gene panel burden tests across our 599 gene subset, with association tests limited to individuals in UKB who had genetically defined European Ancestry due to the well-recognised biases in PGS performance in other ancestries^43^. We controlled for age, sex, recruitment centre and 40 principal components. Variant burden tests were performed using STATA (version 16.0), using linear regression for continuous phenotypes, and logistic regression for binary phenotypes. Associations were tested between individuals with an identified rare variant in any of these DDG2P genes and the remainder of the European UKB population. EA-PGS quintiles were defined using the entire cohort of European UKB. When testing across PGS quantiles, each group was tested against individuals in the middle quintile (i.e. 40-60% EA-PGS) who were not identified as being carriers of likely deleterious rare variants in the DDG2P gene subset. When testing associations within specific types of variants, similarly, the comparison group was those with were not identified as being carriers of likely deleterious variants. When testing smaller subgroups of individuals, those who had previously been identified as putatively deleterious variant carriers were removed from the comparison group.

To define phenotypic “deviators”, we used the highest and lowest scores of fluid intelligence scores (0 and 1 versus 11, 12 and 13), and the top and bottom category for qualifications (no qualifications recorded versus having a degree).

## Data Availability

All data produced in the present study are available upon reasonable request to the authors

## ACKNOWLEDGEMENTS

This research has been conducted using the UK Biobank Resource under Application Number 49847 and 9072. The authors would like to thank Tim Frayling for helpful suggestions, and acknowledge the use of the University of Exeter High-Performance Computing (HPC) facility in carrying out this work. The authors would like to acknowledge support from the University of Exeter, the MRC [MR/T00200X/1] and National Institute for Health and Care Research Exeter Biomedical Research Centre. The views expressed are those of the author(s) and not necessarily those of the MRC, NIHR or the Department of Health and Social Care.

## SUPPLEMENTARY TABLES AND FIGURES

**Figure 1: Trait results across EA-PGS quintiles for variant carriers and non-carriers**

**Table 1: 15 Phenotypic traits tested in UKB**

**Table 2: ICD9 and ICD10 codes used to define clinical phenotypes**

**Table 3: Overall rare variant burden seen in UKB, separated by number of variants seen and type of variants seen**

**Table 4: Association test results for CNV and LoF carriers only (excluding missense variants) for all phenotypes tested in the original group of carriers, by overall variant burden**

**Table 5: Association test results for CNV and LoF carriers only (excluding missense variants) for all phenotypes tested in the original group of carriers per quintile**

**Table 6: Association test results for smaller subsets of the DDG2) gene sets, for all phenotypes tested in the original 599 gene set**

**Table 7: Fluid Intelligence association test results per variant type Table 8: Phenotypic deviator results by variant type**

**Table 9: Association results between clinically diagnosed rare variant carriers and those who do not have a clinical diagnosis but carry a similar rare variant**

