## Supplementary figures and images for "Genetic modifiers of rare variants in monogenic developmental disorder loci"

### Supplementary Figure 1

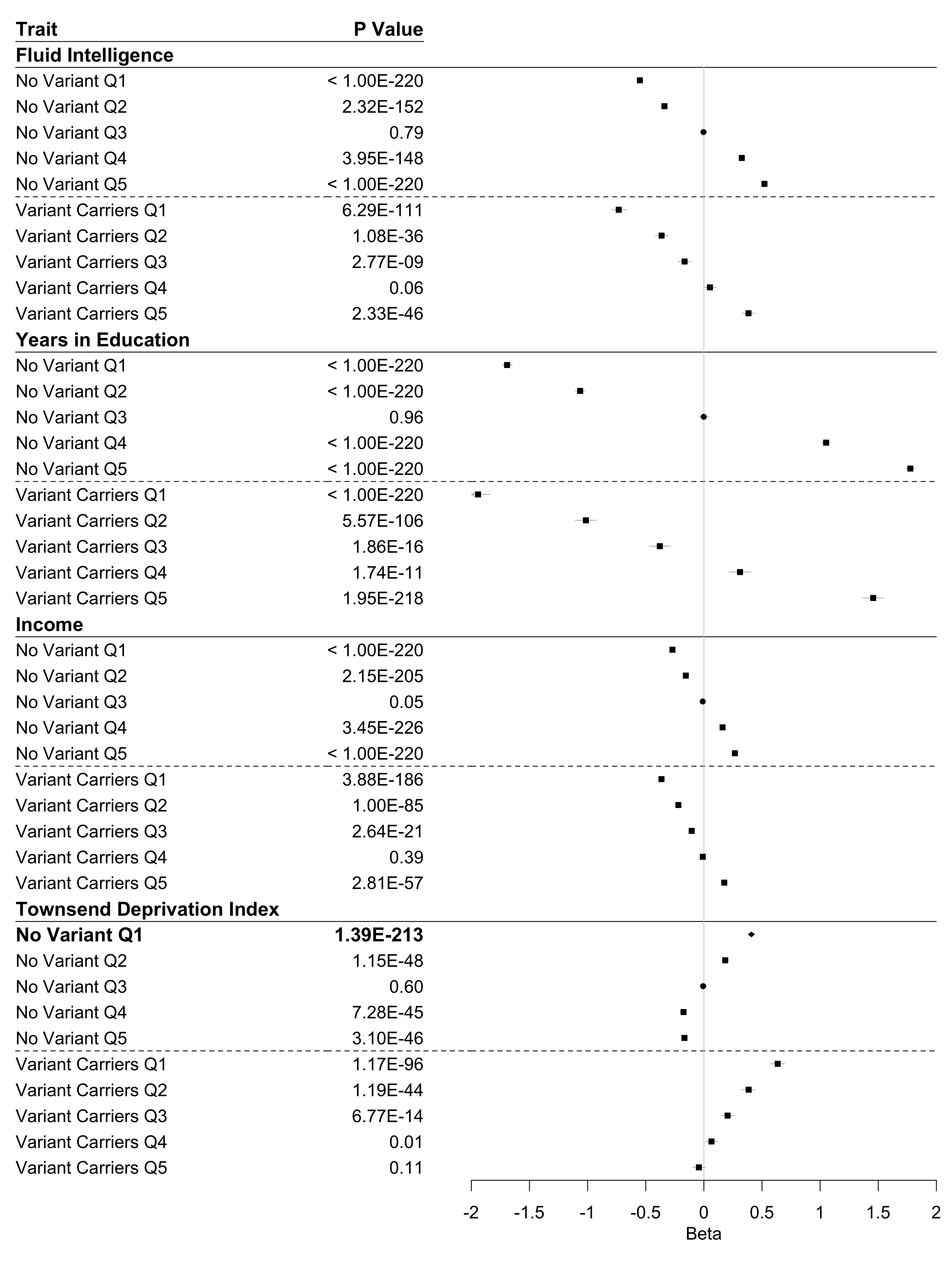
